# A Siamese U-Transformer for change detection on MRI brain for multiple sclerosis, a model development and external validation study

**DOI:** 10.1101/2024.04.05.24305386

**Authors:** Brendan S Kelly, Prateek Mathur, Ronan P Killeen, Aonghus Lawlor

## Abstract

**Background:** Multiple Sclerosis (MS), is a chronic idiopathic demyelinating disorder of the CNS. Imaging plays a central role in diagnosis and monitoring. Monitoring for progression however, can be repetitive for neuroradiologists, and this has led to interest in automated lesion detection. Simultaneously, in the computer science field of Remote Sensing, Change Detection (CD), the identification of change between co-registered images at different times, has been disrupted by the emergence of Vision Transformers. CD offers an alternative to semantic segmentation leveraging the temporal information in the data.

**Methods:** In this retrospective study with external validation we reframe the clinical radiology task of new lesion identification as a CD problem. Consecutive patients who had MRI studies for MS at our institution between 2019 and 2022 were reviewed and those with new lesion(s) were included. External data was obtained from the MSSEG2 challenge and OpenMS. Multiple CD models, and a novel model (NeUFormer), were trained and tested. Results were analysed on both paired slices and at the patient level. Expected Cost (EC) and F2 were independently and prospectively chosen as our primary evaluation metrics. For external data we report DICE and F1 to allow for comparison with existing data. For each test set 1000 bootstrapping simulations were performed by sampling 10 patient samples with replacement giving a non parametric estimate of the confidence interval. Wilcoxon statistics were calculated to test for significance.

**Findings:** 43,440 MR images were included for analysis (21,720 pairs). The internal set comprised of 170 patients (110 for training, 30 for tuning, 30 testing) with 120 females and 50 males, average age of 42 (range 21 – 74). 60 (40 + 20) patients were included for external validation.

In the CD experiments (2D) our proposed NeuFormer model achieved the best (lowest) Expected Cost (EC) (p=0.0095), the best F2 and second best DICE (p<0.0001). At the patient level our NeUFormer model had the joint highest number of True Positive lesions, and lowest number of False negatives (p<0.002). For CD on external data, NeUFormer achieved the highest DICE on both datasets (p<0.0001). NeUFormer had the lowest or joint lowest number of False Positives on external data (p<0.0001 in all cases).

**Interpretation:** Reformulating new lesion identification as a CD problem allows the use of new techniques and methods of evaluation. We introduce a novel Siamese U-Transformer, NeUFormer, which combines concepts from U-Net, Siamese Networks, and vision transformers to create a model with improved small lesion detection and the consistently best EC. Its ability to increase detection of small lesions, balanced with relatively few false positives, and superior generalisability has the potential to greatly impact the field of the identification of radiologic progression of MS with AI.

**Research in context:** *Evidence before this study:* Multiple Sclerosis (MS), a chronic and idiopathic demyelinating disorder of the CNS, is diagnosed using the McDonald criteria based on MRI interpretation. Without a definitive MS biomarker, AI holds promise is for uncovering unique features indicative of MS, improving diagnostics and identifying progression. Research in the field typically centres on segmentation and classification, leaving a gap in evaluating temporal imaging changes. The MSSEG2 challenge has now enabled more research into new lesion identification in MS. Even so, most solutions are based on semantic segmentation architectures and rely on limited metrics for evaluation. The identification of small lesions also remains a challenge. Remote Sensing (RS) is the science of obtaining information about objects or areas from a distance, typically from aircraft or satellites. In the RS literature, Change Detection (CD) refers to the identification of significant alterations in co-registered images captured at different times. In this way CD offers an alternative to semantic segmentation leveraging the temporal information in the data. This field was dominated by convolutional neural networks but has recently been disrupted by transformer-based architectures. Transformers, fuelled by their success in NLP, are gaining popularity across all computer vision tasks due to their larger effective receptive field and enhanced context modelling between image pixels. Inspired by these developments, we incorporate some of these ideas into our NeUFormer model.

*Added value of this study:* This study redefines the task of identifying progression on MRI brain in MS as a CD problem, borrowing concepts from RS. This approach allows for both pixel- and patient-level evaluation and rethinks standard metrics to suit specific clinical needs. This acknowledges the distinction between trivial variation in segmentation and clinically significant change. State-of-the-art CD models are assessed at this task, and a novel model, NeuFormer, is introduced. NeuFormer synergistically combines concepts from the classical U-Net (which was originally intended for brain segmentation), Siamese architecture adaptations specifically for CD, Swin-UNETR (a U-Transformer developed by MONAI to integrate the shifting window structure of the Swin transformer into medical imaging) and ChangeFormer which also uses attention at scale specifically for CD, leveraging improved spaciotemporal reasoning to create a model which is better for small lesion identification and with the consistently lowest EC associated with its decisions.

*Implications of all the available evidence:* Reframing lesion identification as CD enables an alternative to semantic segmentation leveraging the temporal information in the data, enhancing the model’s relevance and customization for specific medical tasks. We also propose the flexible Expected Cost metric, as it facilitates varying action thresholds and helps to customise tools to stakeholder preferences. Siamese vision transformers show promise for CD on MRI in MS including for smaller lesions which are traditionally difficult for computer vision models to identify. This may be to the intrinsic spaciotemporal advantages of vision transformers, with positional embedding, over patch based convolutional methods. NeUFormer’s ability to increase detection of small lesions, balanced with relatively few false positives and excellent generalisability has the potential to greatly impact the field of the identification of radiologic progression of MS with AI.

## Introduction

Multiple Sclerosis (MS), a chronic and idiopathic demyelinating disorder of the CNS, is diagnosed using the McDonald criteria based on MRI interpretation^1,2^. MS differs from many chronic diseases as its imaging features may precede clinical symptoms. As such, imaging plays a central role in diagnosis, tracking progression, and evaluating treatments^1,2^. New T2/FLAIR MS lesions are the primary biomarker for assessing both disease progression and medication response^3,4^. Indeed, lack of new lesions in the CNS is a key indicator of medication effectiveness^5^. Monitoring these lesions, however, is often monotonous and repetitive for neuroradiologists ^6^, and this combined with radiology’s supply-demand challenges^7^ has led to increased interest in methods for automating lesion detection^8^. Over the past two decades, research has heavily focused on computer-assisted segmentation methods ^8^, with a recent surge in AI methodologies^9^. Current research trends are evolving from simple identification of MS lesions on T2/FLAIR to analysing images over different times^8^. The MSSEG2 challenge by The Medical Image Computing and Computer Assisted Intervention Society (MICCAI), targeting new lesion detection, has significantly boosted interest in this research domain^3^. Significant issues remain in the literature with only modest performance reported for new lesion identification^10^ and there are well known difficulties to correctly evaluate stable cases (which outnumber change cases significantly)^11^. Furthermore, identification of smaller lesions remains problematic and is an outstanding issue^10^. The limited kernel size of convolution layers in Fully Convolutional Neural Networks (FCNNs)^12^ can result in sub-optimal performance in modelling long-range spatial information^13^. This can adversely affect the segmentation of lesions of varying sizes^12^.

Remote Sensing (RS) is the science of obtaining information about objects or areas from a distance, typically from aircraft or satellites^14^. In the RS literature, Change Detection (CD) identifies significant alterations in co-registered images captured at different times^15^. CD offers an alternative to semantic segmentation leveraging the temporal information in the data. The type of change varies by application, including alterations in man-made structures, vegetation, and environmental shifts like polar ice cap melting or deforestation. Effective CD models distinguish these changes while filtering out irrelevant variations due to seasonal shifts, shadows, atmospheric changes, and lighting differences^14^. Current leading CD methods primarily use deep convolutional networks (ConvNets) for their strong feature extraction capabilities^13^. Recently, the success of Transformers in Natural Language Processing (NLP) has inspired their application in multiple other domains, and especially in computer vision tasks^14^. Attention-based models such as the original Vision Transformer^16^ and more recently SAM/SAM-Med^17^ and SWIN^18^ continue to improve performance on standard computer vision challenges. This is in part due to their larger effective receptive field than deep ConvNets, enhancing context modelling between image pixels^13^. Indeed, a Siamese vision transformer (ChangeFormer) has reached state of the art performance in CD challenges^14^. There have been some recent medical applications of vision transformers in the medical imaging literature, especially in the domain of semantic segmentation^19,20^. However there are very few works which use these models to leverage the temporal information in these data to enhance the identification and classification of change over time^21,22^. For example the nnU-Net, a leading segmentation model often considered state of the art for MS lesion segmentation, does not use the longitudinal nature of the scans and treats each image individually during training and inference ^23^.

Emerging research demonstrates that “task set-up” or “problem formulation” can have significant impact on the result of medical AI experiments^24^. In this study we reframe the clinical radiology issue of new MS lesion classification on MRI as a CD problem inspired by the RS literature. Here we consider only new or enlarging lesions (as defined by the MAGNIMS criteria^25^) as relevant change. This enables us to consider different evaluation metrics that consider the cost of decisions made by the models rather than just segmentation performance (e.g. DICE score) or lesion identification (e.g. accuracy or F1 score), which are important, but not task specific^26^. The “Expected Cost” (EC) allows individually chosen weights for the error rates (such that missing a lesion can be penalized more) and can be made prevalence-independent. We can evaluate performance on both co-registered slices and the whole 3D stack allowing for scrutiny at both the pixel and patient level. We also describe a novel model for **Ne**w lesion identification, a Siamese **U**-Trans**former**, abbreviated as NeUFormer.

## Methods

### Study design and data sources

This retrospective study with external validation was designed according to both RSNA and ESR published principles ^27,28^ with patient expert involvement ^29^. The EC was independently and prospectively chosen as a suitable primary evaluation metric^26^. The manuscript was prepared using the CLAIM checklist ^30^. We received full IRB approval and the requirement for prospective consent was waived. This research constitutes Level 5A evidence (Data quality and AI model development with external testing) as it represents one retrospective study with internal and external data used for final reporting ^31^. Subsections of this cohort have been published previously but under different experimental conditions with different research questions. The external cohort is publicly available and has been previously described ^3^. All internal data are available from the authors upon reasonable request and the code to recreate these experiments is available on github. Consecutive patients who had at least two MRI brain studies for MS at our institution between January 2019 and December 2022 were reviewed (Figure 1). Those with a new lesion on follow up imaging were included in this study. Images were acquired on a 1.5 T system (SIEMENS MAGNETOM Avanto syngo MR B19, SIEMENS, Munich, Germany). Imaging sequences included a three-dimensional T2 fluid-attenuated inversion-recovery (FLAIR) sequence using the following parameters: acquired voxel size, 1.1 x 1.1 x 1.1 mm; TR 6000 ms; TE 413 ms; TI 2030ms; acquisition time 6 mins 44 s; orientation, sagittal. We used all publicly available data from MSSEG-2^3^ and OpenMS^32^ to externally validate our findings.

**Figure 1:**
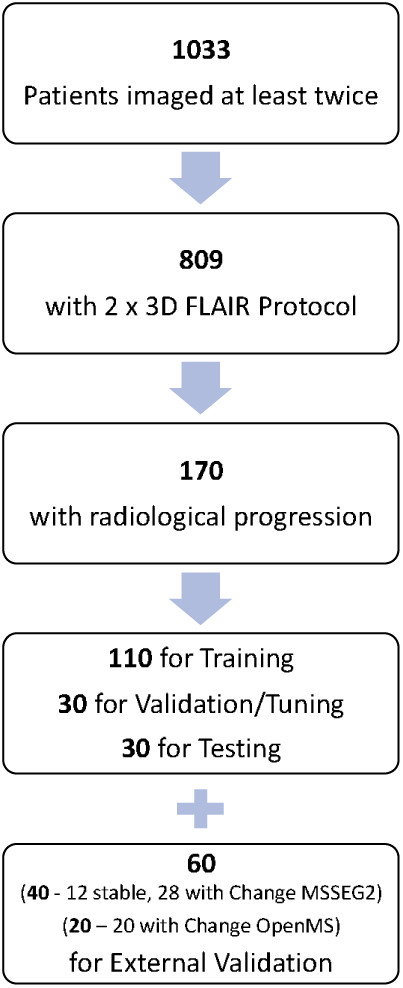
Patient Flowchart. Flowchart of patients included in our study

### Ground truth labelling

A baseline automated segmentation of MS lesions was generated using DeepMedic ^33^. These baseline segmentations were then manually corrected by one of two certified radiologists in their first year post board examination using ITK Snap V3.8.0 ^34^. Cases with progression were initially identified from the radiologic report, and confirmed at a dedicated research re-read. Radiologic progression (new or enlarging lesions) was defined according to the MAGNIMS consensus guidelines^25^. Specifically for new lesions the largest linear measurement for lesion definition had to be 3 mm or more in at least one plane. In the case of enlarging lesions, subtractions of co-registered intensity normalized images were used to confirm that the lesion had unequivocally enlarged. Cases with progression were first segmented and manually corrected as above and then additionally verified by a third radiologist who is a subspecialist neuroradiologist with over 10 years post fellowship experience.

### Image processing

The raw MRI sequences acquired from the scanner were in DICOM format which were anonymized by removing any identifiable information pertaining to the patient or the practitioner. The anonymized DICOM slices were then converted to the NifTI format using the dcm2niix utility (v1.0.20220720). The produced images were then rigidly registered to the first T1 sequence using the FMRIB’s Linear Registration Tool (FLIRT). Rigid body registration was applied with 6 degrees of freedom, no angular search and spline interpolation, rest of the properties were set to their default values. After registration, the FSL Brain Extraction Tool (BET) was applied with a fractional intensity threshold of 0.4 to the first T1 of all patients. This mask was then applied to the remaining scans for each patient respectively. Bias Field correction using the FSL FAST utility was not performed after no contrast improvement was empirically observed. Since the MRI scans in our dataset were acquired from the same Scanner, the need for standardisation was diminished.

Patients with more than two FLAIR images were chosen. Each extracted slice is cropped to the largest brain cross-section while maintaining the image aspect ratio. The cropped slice is then intensity normalised and rescaled to 256x256. A Contrast Limited Histogram Equalization is applied to the cropped slices to enhance the tissue contrast. Slices with no brain volume are discarded. The lesion masks follow a similar pre-processing pipeline except for the Histogram Equalization step. In addition, partial lesions which are too small to be considered progression are removed before the final labels are produced. This is because lesions less than 3mm are not considered to represent progression in the MAGNIMS criteria ^25^ and is standard across similar tasks ^3,11^. The image slices and the difference maps are generated to conform to the usual data structures in CD challenges. Data were partitioned at the patient level into training, validation/tuning and test sets in a ratio of approximately 65:17.5:17.5.

### Model development and training

A novel Siamese U-Transformer (NeUFormer, NeU) was developed (Figure 2). The network is inspired by the “Swin UNETR^12^” semantic segmentation model, and modified to create a Siamese architecture via additional skip connections at each subsampling scale, with a classification head for change detection. The aim here is to enhance the more abstract and less localized information from the later-stage encoded data with spatial details which are learned in the network’s earlier layers. Uniquely for NeUFormer, these skip connections also integrate information from different resolutions. Through this we aim to create a model with improved spaciotemporal awareness, primed for superior change detection.

**Figure 2:**
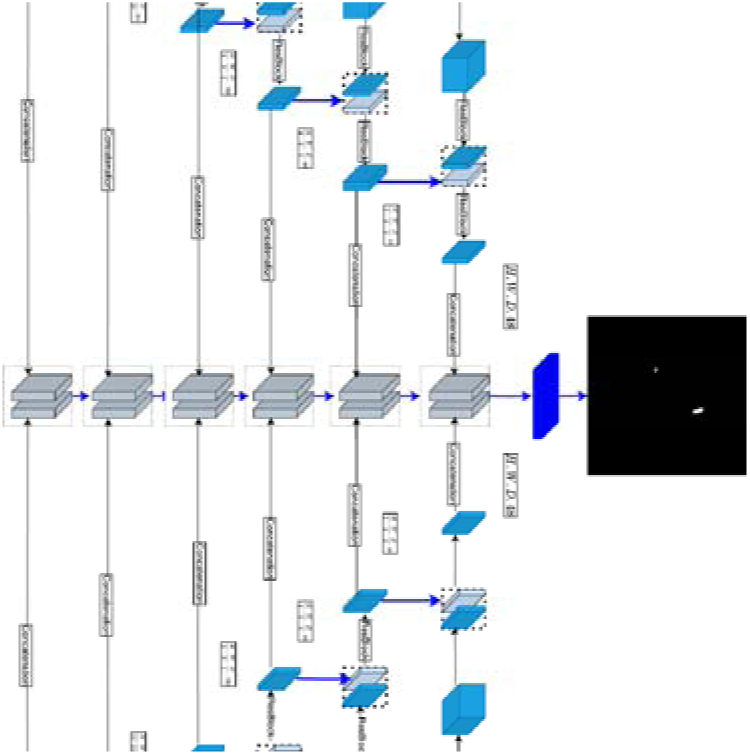
Model Diagram NeUFormer. Diagrammatic representation of the NeUFormer model. (*Navy cuboid = Bottleneck Feature/Head, Light Blue cuboid = Hidden Feature, Navy arrow = Deconvolution, broken line = concatenation*.)

The input to the network is two 256x256 images and the output is a change map. Each arm consists of a “Swin UNETR” encoder (separated into two streams of equal structure with shared weights as in a traditional Siamese network) and decoder. The encoder has 4 stages with 2 transformer blocks at each stage. A modified 2D version of the Swin UNETR 3D shifting window self-attention mechanism is employed, allowing for feature extraction at five different resolutions (Figure 2). For the decoder, NeUFormer also has a U-Shaped FCNN design, with the encoded feature representations used by the decoder at each resolution via skip connections. Extending this concept, the feature representations are concatenated together before and after the encoding part of the architecture via additional “Siamese” skip connections^35^. This is done for each arm and at each resolution, with the goal of producing precise class predictions. We leverage synergy of the Swin transformer’s shifting windows giving hierarchical attention at different scales^18^, the U-Net’s proven ability at biomedical imaging semantic segmentation^36^, and the discriminative power of the Siamese architecture for change detection^35^.

For comparison, baseline models included a fully connected Bitemporal U-Net (BUN) (that concatenates bi-temporal images and passes to a ConvNet to detect changes), two different Siamese U-Nets (which extracts multi-level features of bi-temporal images from a Siamese ConvNet and either concatenates (SUC) them or finds their difference (SUD))^35^, a bitemporal vision transformer (VIT) (which uses a transformer encoder-decoder network to enhance the context-information of ConvNet features via semantic tokens followed by feature differencing to obtain the change map) ^13^, Swin UNeTR (UTR)(which replaces the encoder of a vision transformer with a U-Net) ^12^ and ChangeFormer (CFR) (which utilises a transformer encoder with Multi-Layer Perception decoder in a Siamese network architecture to efficiently render multi-scale long-range details required for accurate CD) ^14^. All models were implemented in PyTorch and trained from scratch (without pretraining) using an NVIDIA GeForce GTX 1080 GPU. Data augmentation was performed with random flip, random re-scale, random crop and Gaussian blur. Models were trained using a combined weighted Cross-Entropy and DICE loss using AdamW optimizer and a batch size of 8. Due to biased classes (only a small % of image was made up by the target (change) class) images with change were used for training along with a matched number of stable controls in a ratio of 1:1.

### Analysis

For the change detection task we completed problem fingerprints for our research questions^26^. Due to the unequal severity of class confusions (with false negatives rated as more important) and an existence based penalization of special outliers the EC (Multi-Class) and F2 score (Per-Class) were chosen prospectively and independently as the primary evaluation metrics for our study^26,37^. EC is a measure of accuracy that can incorporate different disease prevalence and account for differences in impact between false positives and false negative predictions. The closer to 0 the better the expected cost; while scores of >1 are possible, the indicate a futile model. Following a stakeholder focus group a cost matrix of [[0.0, 10.0], [1.0, 0.0]] was used to calculate the EC ^29^. This penalises FNs 10 times higher than FPs. We also report the precision, recall, F0.5, F1 and DICE scores to allow for comparison with other studies. For the DICE, a score of 1.0 was recorded both where a model correctly predicted the positive change map or a blank change mask as many cases have “no change”. We also evaluate on small lesions (those with segmentation masks with only connected components less than 100 pixels) separately as this is a consistently a problem in the literature. The 100 pixel connected component limit was chosen as this is the limit at which segmentation models have been shown to deteriorate for 256x256 images^17^. For each test set 1000 bootstrapping simulations were created by sampling 10 patient samples with replacement giving a non parametric estimate of the confidence interval. Wilcoxon statistics were calculated to test for significance. To minimise hypothesis testing only the best and second best model were compared for primary evaluation metrics (EC, F2, F1, DICE).

We also evaluate our models on external data (MSSEG-2^3^ and OpenMS^32^) to test their generalisability and potential clinical applicability. For both cases only the training data could be accessed, and it was then processed according to the same pipeline as the internal data. The mix of cases that are positive and negative for progression in the external data allows us to report accuracy, sensitivity and specificity at the patient level (3D). Total True Positive, True Negative and False Positive lesions are reported for all datasets.

## Results

In total 43,440 MR images were included for analysis (21,720 pairs including train, tuning, test and external data). The internal set comprised of 170 patients (110 used only for training, 30 for tuning and 30 testing) and the external set comprised of 60 patients for external validation (40 from MSSEG2 and 20 from OpenMS) (Figure 1). There were 120 females and 50 males in the internal data with and average age of 42 (range 21 – 74). Demographics are summarised in Table 1. Due to the nature of the condition, the target (change) class was underrepresented in the images with the target class representing only 0.0001% of the total pixels (see Table 2).

**Table 1.** Patient Demographics.

| Total Participants | Average Age |  | Age Range |
| --- | --- | --- | --- |
| 170 |  |  |  |
|  | 42.3 | Minimum | 21 |
|  |  | Maximum | 74 |
| Gender | Gender Count | Change Instances |  |
| Male | 50 | 212 |  |
| Female | 120 |  |  |
| MRI Studies Range | MRI Studies | Total MRI Studies | MRI Studies Average |
| Min | 2 | 496 |  |
| Max | 5 |  | 2.9 |

**Table 2.** Prevalence of the change class (progression) in the internal test and external data.

| Test Set Lesions | Patients | Images (256x256) | Number positive images | Number Positive Pixels (%) | Lesion dimension (pixels): Mean, StD (range) |
| --- | --- | --- | --- | --- | --- |
| Internal | 30 | 6081 | 437 | 43823 (0.000109%) | 164.71, 111.42 (14 - 407) |
| MSSEG2 | 28 (of 40) | 7361 | 629 | 53408 (0.00011%) | 53.95, 52.44, (6 - 360) |

For the two primary evaluation metrics, NeuFormer achieved the best (lowest) EC of 0.467 (p=0.0095) and had the highest F2 score 0.329 (Figure 3). NeuFormer also achieved the lowest EC and F2 for small lesions (lesions with connected components less than 100 pixels). Results of the CD experiments are summarised in Table 3A for all lesions and Table 3B for small lesions only. Additionally Figure 4 shows the qualitative difference in output for the different models. For patient level evaluation, NeUFormer had the joint highest number of True Positive lesions (p=0.0011) and lowest number of False negatives (p<0.0001) (Table 4).

**Figure 3:**
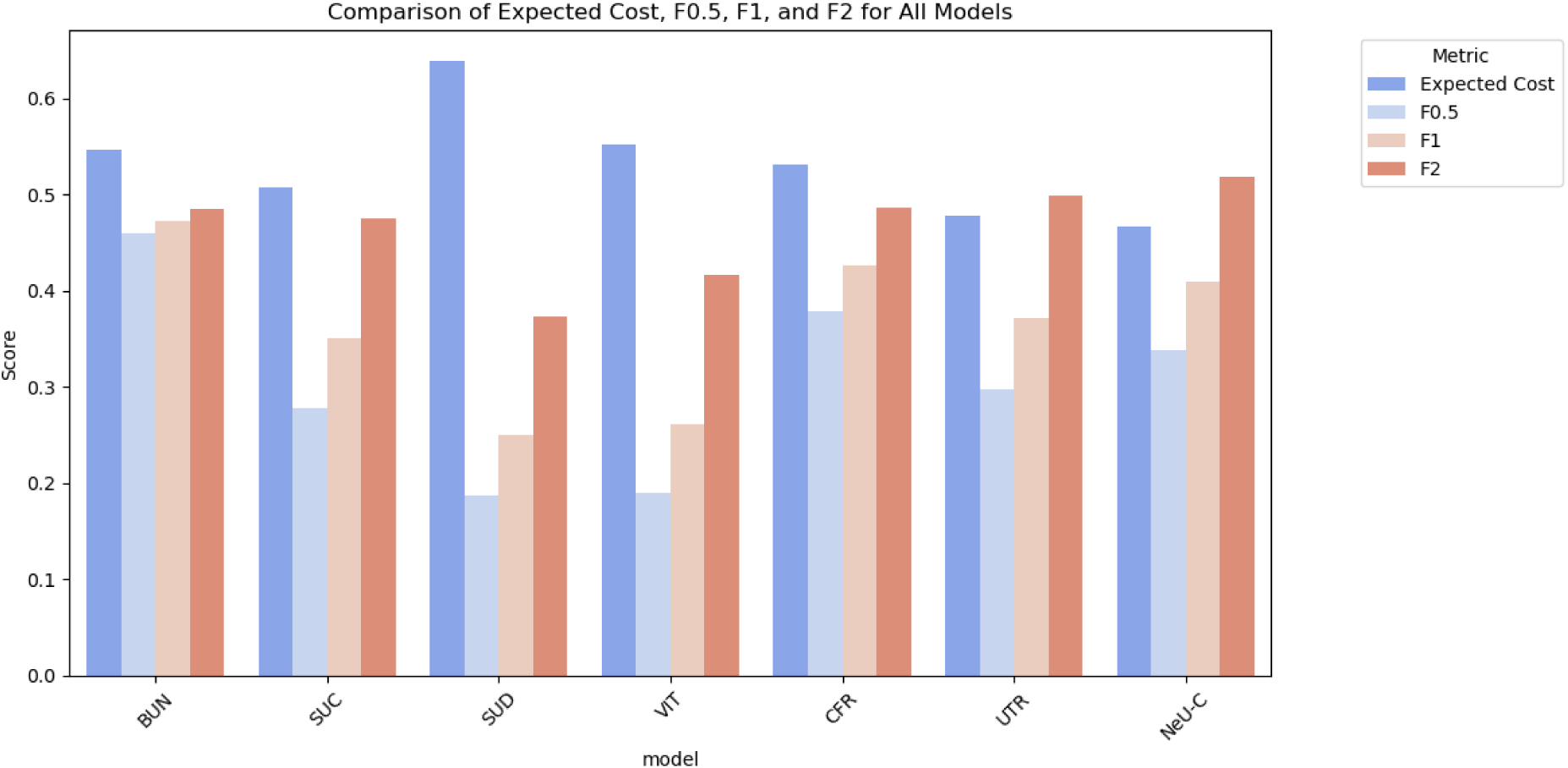
Quantitative Results 2D. Comparison of the performance metrics for all models on the internal data (2D). Bitemporal U-Net (BUN), Siamese U-Net with Concatenation (SUC), with Difference (SUD), bitemporal Vision Transformer (VIT), ChangeFormer (CFR), Swin UNeTR (UTR), NeUformer (NeU).

**Figure 4:**
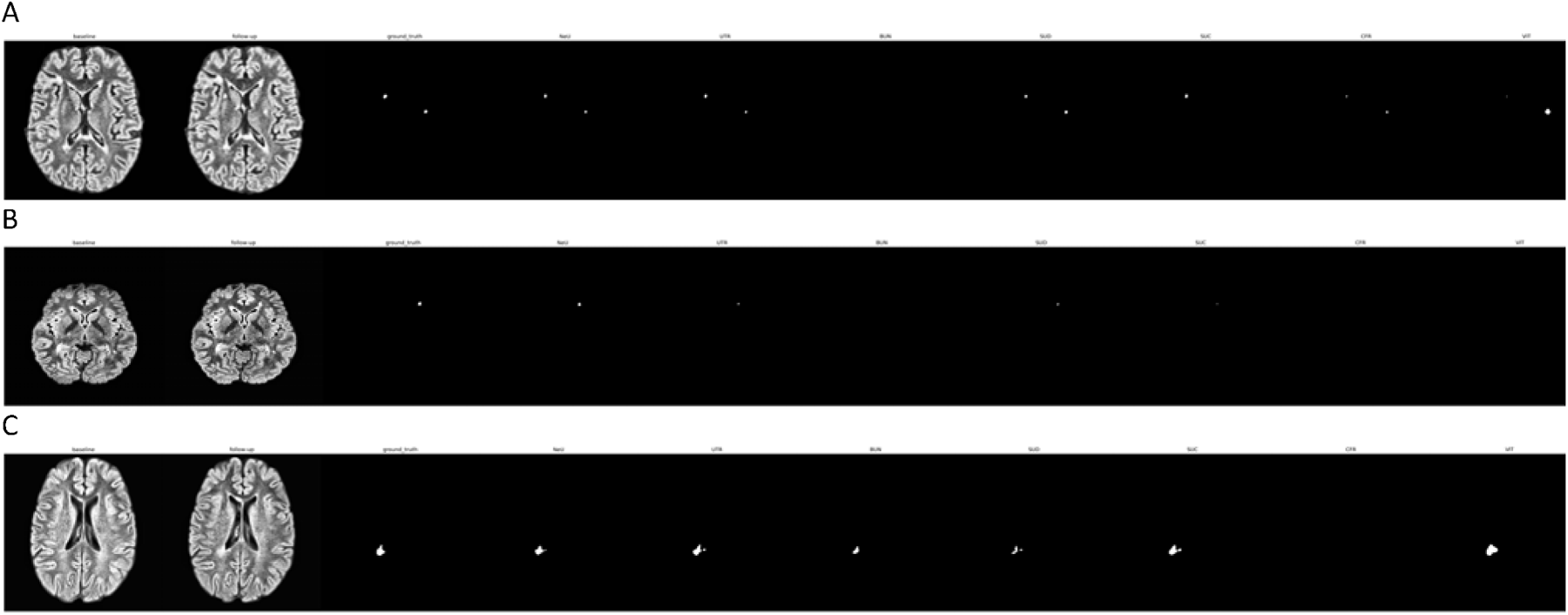
Qualitative Results. Qualitative comparison of different CD methods on the internal data (A), MSSEG2 (B) and OpenMS (C). Bitemporal U-Net (BUN), Siamese U-Net with Concatenation (SUC), with Difference (SUD), bitemporal Vision Transformer (VIT), ChangeFormer (CFR), Swin UNeTR (UTR), NeUformer (NeU).

**Table 3A.** CD Results internal data, all lesions.

| Model | Expected Cost | DICE | Precision | Recall | F0.5 | F1 | F2 |
| --- | --- | --- | --- | --- | --- | --- | --- |
| BUN | 0.546 | **0.932\*** | **0.451** | 0.494 | **0.459** | **0.472\*** | 0.485 |
| SUC | **0.507** | 0.886 | 0.244 | 0.624 | 0.277 | 0.350 | 0.476 |
| SUD | 0.638 | 0.848 | 0.161 | 0.559 | 0.187 | 0.249 | 0.374 |
| VIT | 0.551 | 0.873 | 0.161 | **0.693** | 0.190 | 0.261 | 0.417 |
| CFR | 0.531 | **0.887** | **0.353** | 0.535 | **0.379** | **0.426** | **0.485** |
| UTR | **0.478** | 0.851 | 0.262 | **0.644** | 0.297 | 0.372 | **0.498** |
| NeU-C | **0.467\*** | **0.915** | **0.302** | **0.632** | **0.337** | **0.409** | **0.519\*** |

Red is best, blue second best and bold third best
Bitemporal U-Net (BUN), Siamese U-Net with Concatenation (SUC), with Difference (SUD), bitemporal Vision Transformer (VIT), ChangeFormer (CFR), Swin UNeTR (UTR), NeUformer (NeU).
\*denotes statistical significance

**Table 3B.** CD Results internal data, all lesions.

| Model | Expected Cost | DICE | Precision | Recall | F0.5 | F1 | F2 |
| --- | --- | --- | --- | --- | --- | --- | --- |
| BUN | 0.666 | **0.940\*** | **0.181** | 0.363 | **0.201** | **0.241\*** | **0.302** |
| SUC | **0.621** | **0.894** | 0.081 | 0.475 | 0.097 | 0.139 | 0.241 |
| SUD | 0.670 | 0.855 | 0.049 | 0.503 | 0.060 | 0.089 | 0.176 |
| VIT | 0.663 | 0.881 | 0.048 | **0.519** | 0.059 | 0.088 | 0.176 |
| CFR | 0.665 | 0.893 | **0.118** | 0.386 | **0.137** | 0.181 | 0.265 |
| UTR | **0.527** | 0.858 | 0.114 | **0.549** | 0.135 | **0.189** | **0.311** |
| NeU-C | **0.516\*** | **0.922** | **0.125** | **0.552** | **0.148** | **0.205** | **0.329\*** |

Red is best, blue second best and bold third best
Bitemporal U-Net (BUN), Siamese U-Net with Concatenation (SUC), with Difference (SUD), bitemporal Vision Transformer (VIT), ChangeFormer (CFR), Swin UNeTR (UTR), NeUformer (NeU).
\*denotes statistical significance

**Table 4.**
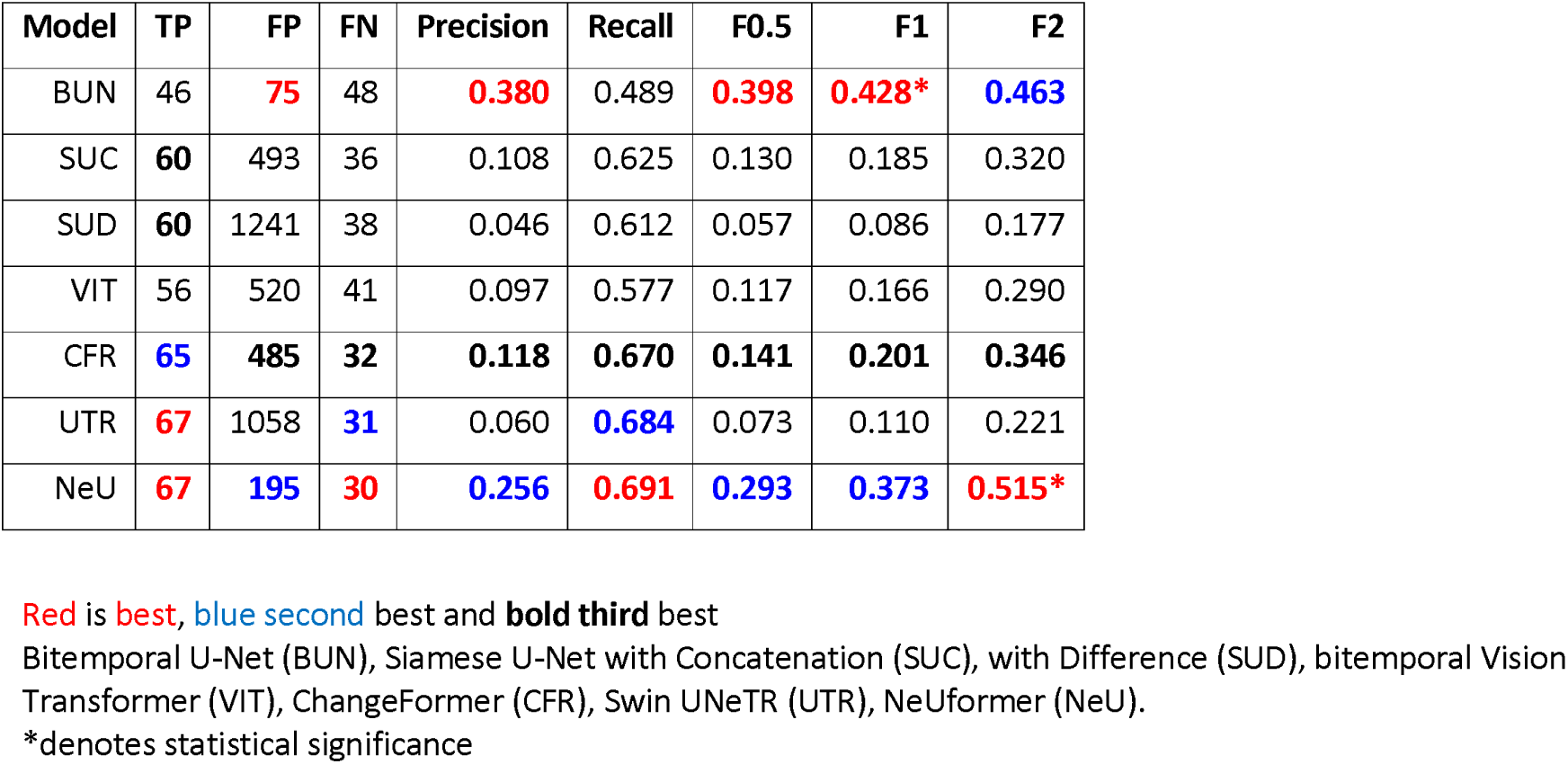
Patient level results internal data.

For external validation we focus on the widely used metrics of DICE and F1 to allow for comparison. Two external datasets MSSEG2 and OpenMS were analysed. MSSEG2 has both progression and stable cases allowing for analysis of progression identification at the patient level. NeUFormer consistently performed well across both datasets, achieving the highest DICE on both (p<0.0001), the second highest F1 for OpenMS, and third highest F1 for MSSEG2 (Table 5 A and B). For patient level evaluation on external data the highest progression accuracy was achieved by UTR for the MSSEG2 dataset but the model had 723 False Positives (compared to 22 by the best performing model NeUFormer) with a specificity of 0.0 for progression Table 6.

**Table 5A.** A External CD Results (MSSEG-2)

| Model | Expected Cost | DICE | Precision | Recall | F0.5 | F1 | F2 |
| --- | --- | --- | --- | --- | --- | --- | --- |
| BUN | 0.527 | **0.932** | **0.711** | 0.486 | **0.650** | **0.577** | 0.518 |
| SUC | **0.413** | 0.928 | 0.520 | **0.624** | 0.538 | 0.567 | **0.600** |
| SUD | **0.375\*** | 0.914 | 0.452 | **0.678** | 0.485 | 0.543 | **0.617** |
| VIT | 0.499 | 0.924 | 0.408 | 0.554 | 0.431 | 0.470 | 0.517 |
| CFR | 0.615 | **0.928** | **0.810** | 0.391 | **0.667** | 0.527 | 0.436 |
| UTR | **0.388** | 0.871 | 0.532 | **0.649** | 0.552 | **0.585\*** | **0.622\*** |
| NeU | 0.549 | **0.941\*** | **0.747** | 0.461 | **0.665** | **0.570** | 0.500 |

Red is best, blue second best and bold third best
Bitemporal U-Net (BUN), Siamese U-Net with Concatenation (SUC), with Difference (SUD), bitemporal Vision Transformer (VIT), ChangeFormer (CFR), Swin UNeTR (UTR), NeUformer (NeU).
\*denotes statistical significance

**Table 5B.**
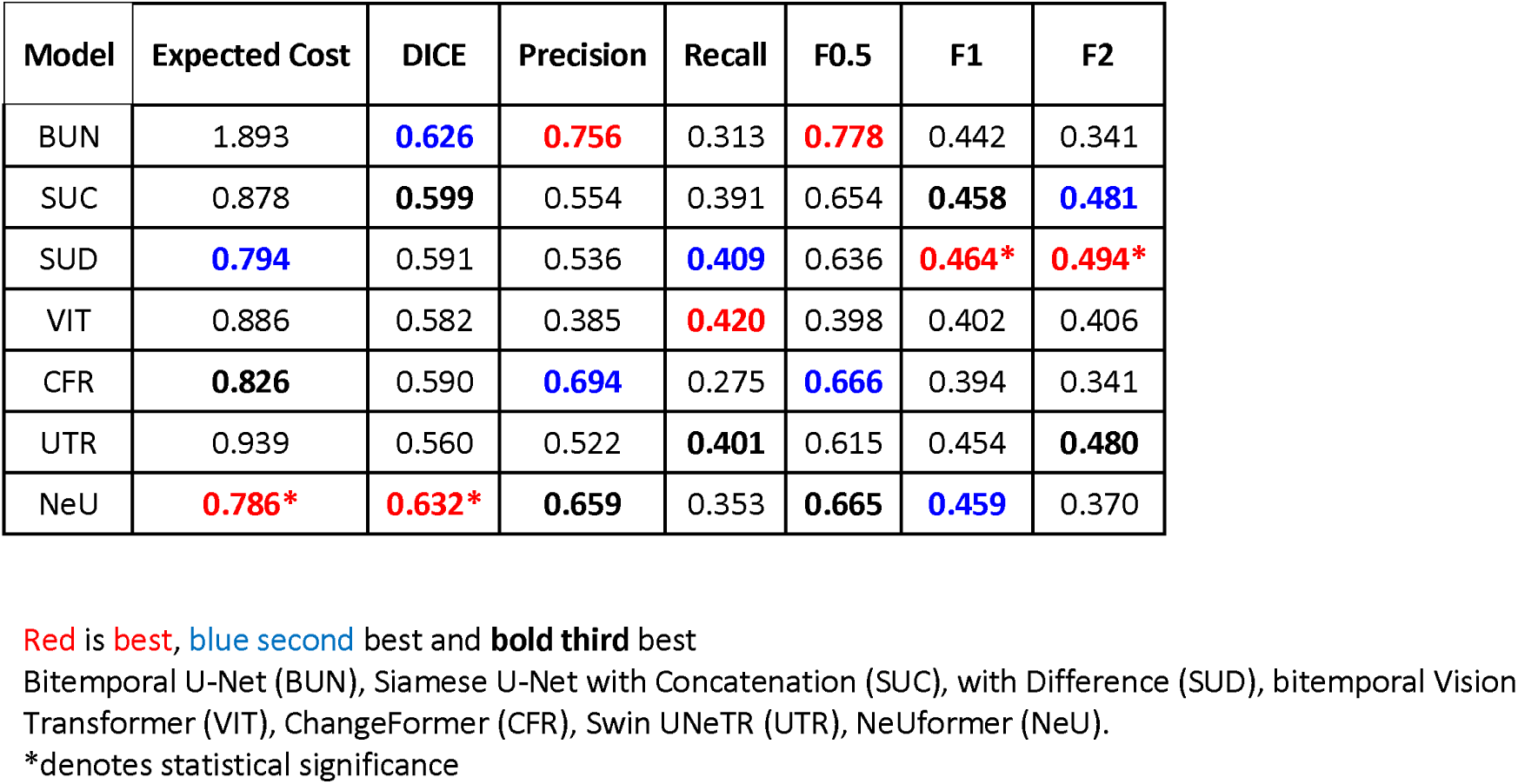
External CD Results (OpenMS)

| Model | Expected Cost | DICE | Precision | Recall | F0.5 | F1 | F2 |
| --- | --- | --- | --- | --- | --- | --- | --- |
| BUN | 1.893 | **0.626** | **0.756** | 0.313 | **0.778** | 0.442 | 0.341 |
| SUC | 0.878 | **0.599** | 0.554 | 0.391 | 0.654 | **0.458** | **0.481** |
| SUD | **0.794** | 0.591 | 0.536 | **0.409** | 0.636 | **0.464\*** | **0.494\*** |
| VIT | 0.886 | 0.582 | 0.385 | **0.420** | 0.398 | 0.402 | 0.406 |
| CFR | **0.826** | 0.590 | **0.694** | 0.275 | **0.666** | 0.394 | 0.341 |
| UTR | 0.939 | 0.560 | 0.522 | **0.401** | 0.615 | 0.454 | **0.480** |
| NeU | **0.786\*** | **0.632\*** | **0.659** | 0.353 | **0.665** | **0.459** | 0.370 |

Red is best, blue second best and bold third best
Bitemporal U-Net (BUN), Siamese U-Net with Concatenation (SUC), with Difference (SUD), bitemporal Vision Transformer (VIT), ChangeFormer (CFR), Swin UNeTR (UTR), NeUformer (NeU).
\*denotes statistical significance

**Table 6.** Patient level results MSSEG-2.

| Model | TP | FP | FN | Precision Progression | Recall Progression | Specificity Progression | Accuracy Progression |
| --- | --- | --- | --- | --- | --- | --- | --- |
| BUN | 44 | **39** | 93 | 0.451 | **0.793** | 0.500 | 0.821 |
| SUC | **56** | 88 | **84** | **0.491** | 0.771 | **0.667** | **0.964** |
| SUD | **56** | 192 | **81** | **0.481** | **0.839** | 0.417 | **0.929** |
| VIT | 40 | 79 | 93 | 0.462 | 0.727 | **0.750** | 0.857 |
| CFR | 43 | **66** | 96 | 0.440 | 0.688 | **0.833** | 0.786 |
| UTR | **74** | 723 | **71** | **0.500** | **1.000** | 0.000 | **1.000** |
| NeU-C | **48** | **22** | 92 | **0.481** | 0.743 | **0.750** | **0.929** |

Red is best, blue second best and bold third best
Bitemporal U-Net (BUN), Siamese U-Net with Concatenation (SUC), with Difference (SUD), bitemporal Vision Transformer (VIT), ChangeFormer (CFR), Swin UNeTR (UTR), NeUformer (NeU).

## Discussion

In this study we reformulate the problem of diagnosis progression on MRI brain in MS into a CD problem inspired by the RS literature. This allows us to evaluate model performance at both the pixel and patient level, and rethink the existing evaluation methods, choosing metrics relevant to the specific problem space. In this way, we can consider which model has the lowest clinical cost associated with its decisions rather than just an evaluation of its segmentation performance. This acknowledges the distinction between trivial variation in segmentation and clinically significant change. We evaluate state of the art CD models and we introduce a novel model, NeuFormer, which synergistically combines concepts from the classical U-Net, Siamese architectures, and Vision Transformers with Shifting windows to create a model with the consistently lowest EC associated with its decisions. In particular, it also has the lowest cost and highest recall and F2 performance when examining small lesions, and demonstrates robust generalisability to external data. Its ability to increase detection of small lesions, balanced with relatively few false positives, has the potential to greatly impact the field of the identification of radiologic progression of MS with AI. Our two primary evaluation metrics were expected cost and F2 score. These metrics emphasise the importance of minimising False Negatives as in our use case this could mean missing an opportunity to change or initiate a treatment. We also consider F1 and DICE, especially for the external data, as these metrics are more commonly used in the literature related to the those datasets.

In our internal dataset, NeuFormer performs best in terms of EC and F2, for both all lesions and small lesions only. Its F1, F0.5 and precision all showed a relative increase in performance for small lesions showing that in addition to its ability for find small lesions (high recall), it also had relatively fewer false positives (better precision). The generalisability of NeUFormer is clear from its performance on OpenMS and on MSSEG2 achieving the best DICE score for both datasets. It has only the third best F1 on MSSEG2 (after UTR and BUN) and second best on OpenMS (after SUD). This shows good performance across the datasets, not dropping out of the top 3 despite the heterogenous data. UTR achieved the best F1 on MSSEG2, however it predicted change on every slice, rendering it useless clinically. When we consider the results at a patient level, our model had both the highest number of TPs and the lowest FPs on internal data. For MSSEG-2 NeUFormer again had the lowest number of False positives, and the third highest TPs (after UTR and SUD). However again due to UTRs high number of FPs (723) it is not usable functionally as a clinical decision model. The other models with comparable performance SUC and SUD also had higher FPs (88 and 192 respectively). This was also borne out in the second external dataset OpenMS where again our model had the fewest FPs.

We believe our models performance and generalisability are in part due to the intrinsic advantages of vision transformers in terms of spaciotemporal awareness, especially when synergistically combined with a U-Net inspired encoder module tried and tested in the medical image domain. Then our novel model design which combines the sensitive UTR encoder with the intrinsically discriminative Siamese architecture provides a balance of consistently identifying clinically relevant changes without excessive FPs. Trivial changes in semantic segmentations are often attributable to differences in inter- and intra-user variability, the CD paradigm allows us to focus on these relevant changes when evaluating the models. In this context the importance of the NeUFormer’s discriminative ability becomes clear, allowing the identification of progression at the patient level without overburdening a decision system with FPs.

Our study has several limitations. The retrospective study design limits the level of evidence. Furthermore, as our internal experiments only involved those patients with progression, there is a selection bias. While this is a common issue in clinical radiology research^38^ it remains a clear limitation. Another key issue was the class imbalance. The target class is very underrepresented in the problem of change detection in MS. Indeed in our datasets only 0.0001% of the pixels were in the target class. For this reason it is necessary to oversample the patients with change in the training process. Even with our oversampling method, we still need to use a Cross Entropy loss weighted 1000:1 in favour of the change class to get the model to make any predictions of change. Since the models are agnostic to the nature of the input images, the slice wise approach provides samples of both cases with and without change. A patient-level control approach will only add to an already imbalanced dataset, making the problem of under representation of the target class worse. We then include stable patients in the evaluation to ensure the model works across different groups.

Our sample size was modest, but 170 pairs with change for training compares favourably to MSSEG2 (the largest existing dataset) which comprised only 100 pairs of patients in total, of whom half were stable. Images were resized to 256x256 and primarily assessed in 2D. While there is loss of the 3D information, our model does integrate the temporal information intrinsic to the interpretation task. Due to positional embedding the transformer based methods can interrogate the whole slice rather than subsections. The alternative models are mostly patch based and thus do not take the whole 3D volume en-bloc. In this way a trade-off is necessary at one level of abstraction or another. The demographic information for internal and external data does not contain information on ethnicity, meaning subgroup analysis was not possible. While the internal data was acquired at one institution on one MRI scanner, the external data was heterogenous with different nations, hospitals, protocols, scanners and even a mix of 1.5 and 3 Tesla magnets.

Prospective evaluation would be necessary before our model could be implemented into clinical practise. The robust external validation results show that the method holds promise but practical implementation remains a challenge. Due to ethical considerations and patient preference an autonomous change detection model it is not currently feasible or desirable^29^. Therefore we propose that NeUFormer could be used to screen for studies likely to contain significant change and be used to triage radiologist workflow. As the output of the model is a binary change map (Figure 4), an intuitive explanation is given to the interpreting physician. Furthermore scalability and computational performance is less of an acute issue than in use cases such as stroke as the proposed use case refers mainly to outpatient imaging. Our approach holds further potential clinical utility outside of MS, in fields where monitoring for change is crucial, such as oncologic imaging, treatment response evaluation, and screening programs.

In summary, reformulating new lesion identification as a CD problem allows the use of new techniques and methods of evaluation. We expect this format to be used to drive new innovations in AI in MS imaging, and that the experimental design can be easily applied to other diseases and modalities. Our novel NeUFormer model combines concepts from U-Net, Siamese Networks, and vision transformers to create a model with the consistently lowest cost associated with its decisions, including for smaller lesions and has the potential to screen for progression of MS on MRI brain.

## Data Availability

All data produced in the present study are available upon reasonable request to the authors

